# Autoantibodies neutralizing type I interferons in patients with life-threatening COVID-19 pneumonia: a meta-analysis from 2020–2026

**DOI:** 10.64898/2026.08.06.26359907

**Authors:** Elsa Feredj, Qian Zhang, Paul Bastard, Jean-Laurent Casanova, Aurélie Cobat

## Abstract

Autoantibodies neutralizing type I IFNs (AAN–IFN-I) have been found in significant proportions of cases of severe, critical, and fatal COVID-19 pneumonia. We performed a systematic review of 54 studies reporting auto-Abs against type I IFNs and a meta-analysis of 20 studies reporting auto-Abs neutralizing type I IFNs published between 2020 and 2026. The meta-analysis included data for 11,380 SARS-CoV-2-infected individuals from Europe, North America, South America, Asia, the Middle East, North Africa and international multicenter cohorts, including 7,814 with severe or critical disease (69%). The pooled prevalence of AAN–IFN-I was estimated at 7.9% (95% CI, 6.0–10.4). Disease severity was strongly associated with AAN–IFN-I prevalence (OR, 11.7; 95% CI, 7.6–17.9; *P*=5×10⁻²⁹). The pooled prevalence of AAN-IFN-I reached 11.4% (95% CI, 10.2– 12.7%) in patients with severe or critical COVID-19 and 15.3% (95% CI, 12.1–19.2%) in those who died. The prevalence of AAN–IFN-I increased with age in patients with severe, critical, or fatal COVID-19. AAN-IFN-I probably accounted for about 1.1 million of the 7.1 million deaths from COVID-19. AAN-IFN-I are strong, common, global determinants of life-threatening COVID-19 pneumonia.

## Introduction

Marked interindividual clinical heterogeneity was observed among unvaccinated individuals infected with pre-Omicron SARS-CoV-2 variants. About 20% developed pneumonia, with ∼3% progressing to critical respiratory failure(1, 2), with an infection fatality rate of ∼1%. Age is a major determinant of outcome(3, 4), with mortality increasing exponentially and reaching ∼10% in individuals over 85 years of age(4, 5). A key advance in our understanding of this heterogeneity came with the identification of inborn errors of immunity (IEI) affecting the type I interferon (IFN-I) pathway in patients with life-threatening COVID-19 pneumonia, highlighting the essential role of IFN-I in antiviral defense(2, 6, 7). Rare monogenic defects impairing IFN-I induction or signaling — affecting TLR3(2, 8), TLR7(9, 10), IFIH1(11), TBK1(12), TRIF, IRF3, UNC93B1(8), IRF7(8, 13), IRF9(8), IFNAR1(14, 15), IFNAR2(16), STAT2(17), TYK2(18), IRAK4, or MYD88(19) — are found in ∼1–5% of patients with critical disease under the age of 60 years(9, 10, 20), and ∼10% of children with severe COVID-19 pneumonia(21). These IEI confer susceptibility to other severe viral infections, such as herpes simplex encephalitis or critical influenza, in otherwise healthy individuals and display incomplete penetrance. Indeed, most individuals were free of severe viral infections before developing life-threatening COVID-19. X-linked TLR7 deficiency is the only IEI reported exclusively in patients with severe COVID-19 pneumonia to date(9, 10, 22, 22–25).

The discovery of an autoimmune phenocopy of IEI affecting IFN-I immunity and underlying life- threatening COVID-19 was first reported in 2020(7). Auto-Abs against IFN-I were first described more than 40 years earlier, but they were largely considered clinically silent. During the first wave of COVID-19, pre-existing auto-Abs neutralizing IFN-α2 and/or IFN-ω were identified and shown to account for ∼15% of critical cases and ∼20% of COVID-19-related deaths in two international studies spanning over 40 countries and including thousands of individuals(5, 7, 26). About 10% of unvaccinated children hospitalized for COVID-19 pneumonia harbor such auto-Abs (21). These auto-Abs are also present in the general population, with a prevalence of ∼1% in individuals below the age of 70 years and >4% in those over the age of 70 years(5, 27). Following the initial 2020 report, the presence of neutralizing auto-Abs against type I IFNs in patients with COVID-19 was investigated in multiple independent cohorts worldwide, and shown to underlie other severe viral diseases(28). Previous systematic reviews and meta-analyses have investigated anti-type I IFN auto-Abs in COVID-19(28–32). This study provides a comprehensive review of 54 studies and a meta-analysis restricted to functionally neutralizing auto-Abs. We obtained stratified data directly from the corresponding authors, making it possible to perform analyses by disease severity, age, and mortality.

## Methods

### Search strategy

We performed a systematic literature search to identify original studies investigating AAN–IFN-I in patients with COVID-19. We systematically screened all articles citing the initial 2020 report with PubMed and then performed a systematic search in PubMed based on combinations of the following keywords: (“COVID-19” OR “SARS-CoV-2”) AND (“autoantibodies” OR “auto- antibodies”) AND (“interferon” OR “IFN”) AND (“patients” OR “cohort” OR “clinical” OR “case series”). The search was limited to studies published between March 2020 and July 2026. We excluded reviews, meta-analyses, editorials, comments, non-human studies, and studies not reporting original data. Titles and abstracts were screened, and a full text review was then performed on the potentially relevant studies. Original studies reporting primary data for the prevalence and/or functional characterization of AAN–IFN-I in patients with confirmed SARS-CoV-2 infection were assessed for eligibility.

### Selection criteria

Studies were considered eligible if they reported: (1) diagnostic methods for SARS-CoV-2 infection; (2) clinical classifications of COVID-19 severity; (3) methods used for the detection of auto-Abs against type I; (4) vaccination status; and (5) SARS-CoV-2 variants.

#### (1) Diagnostic methods for SARS-CoV-2 infection

We included studies in which SARS-CoV-2 infection was confirmed by RT-PCR on respiratory specimens (nasal or nasopharyngeal swabs, or other respiratory samples), with or without additional modes of diagnosis (chest CT imaging or serology). Serological studies were considered eligible only if the findings were supported by a compatible clinical presentation. Studies in which the diagnostic method was not specified were excluded from analyses requiring the confirmation of infection status.

#### (2) Clinical classification of disease severity

The severity of SARS-CoV-2 infection was classified according to clinical presentation and the level of care required. Patients were classified as having asymptomatic/mild (no pneumonia and managed as outpatients), moderate (hospitalized for pneumonia but not requiring oxygen), severe (requiring <6 L/min supplemental oxygen), or critical (requiring high-level respiratory support— CPAP, BIPAP, high-flow oxygen, or intubation—experiencing septic shock or requiring ICU admission for organ dysfunction) disease. We grouped together asymptomatic/mild/moderate cases and severe/critical cases. Some of the included studies also evaluated individuals from the general population without SARS-CoV-2 infection.

#### (3) Detection of Auto-Abs against Type I IFN (type I IFNs)

Two approaches were used to detect auto-Abs against type I interferons (IFN-I): binding assays and functional neutralization assays. Binding assays detect antibody binding without providing functional evidence of neutralization, whereas neutralizing activity is assessed in cell-based assays measuring the ability of auto-Abs to inhibit IFN-I–induced signaling or antiviral activity.

#### (4) Vaccination Status

Studies were included if they (i) explicitly reported vaccination status with extractable numbers of vaccinated and unvaccinated patients, (ii) enrolled exclusively unvaccinated individuals, or (iii) included patients recruited before the start of large-scale COVID-19 vaccination in the country concerned, defined as December 2020. Studies performed after this date were included only if vaccination status was clearly specified or if less than 10% of the general population of the country concerned was fully vaccinated at the end of the inclusion period. Studies with unclear inclusion periods (e.g., broadly reported as “2021” without specification of timing relative to vaccine rollout) or without explicit vaccination data were excluded.

#### (5) SARS-CoV-2 Variants

Similarly, given the substantially different clinical spectrum and risk profile of severe and critical COVID-19 during the Omicron wave, we excluded studies including patients infected during periods compatible with circulation of the Omicron variant. The World Health Organization designated Omicron (B.1.1.529) as a Variant of Concern in November 2021, and it spread rapidly worldwide during December 2021. As the infecting strain was rarely specified, variant attribution was inferred from the reported inclusion period in each country. Studies were included up to December 2021, and those with recruitment extending beyond this period or without precise inclusion dates were excluded.

### Data analysis

For the narrative review, we aimed to provide an overview of all studies investigating AAN–IFN- I in patients with SARS-CoV-2 infection across independent cohorts worldwide, regardless of the methods used for the diagnosis of SARS-CoV-2 infection, disease severity classification, autoantibody detection, vaccination status, or circulating SARS-CoV-2 variants. For the meta-analysis, only pre-Omicron studies conducted in predominantly unvaccinated individuals, with defined severity criteria and functional assessments of the neutralizing activity of auto-Abs, were included.

Overall pooled prevalence estimates and the corresponding 95% confidence intervals (95% CI) were obtained with random-effects models fitted by restricted maximum likelihood (REML) to logit-transformed individual prevalence estimates with the metafor R package. A continuity correction of 0.5 was applied to studies with zero events. Between-study heterogeneity was assessed with Cochran’s Q test, the I^2^ statistic, and the between-study variance (τ^2^). Meta- regression analyses were conducted to evaluate the effects of age, sex, disease severity, and neutralizing activity on prevalence estimates. Subgroup analyses were performed for disease severity, age category, neutralization assay strategy, and mortality. Sensitivity analyses excluding the study by Bastard et al. (2021) were also conducted.

### Data extraction and quality assessment

Two reviewers independently screened the titles and abstracts of all identified studies, excluding those irrelevant to the research topic. The full texts of the remaining papers were retrieved and further evaluated by the reviewers, with the exclusion of studies failing to meet the inclusion criteria. Disagreements were resolved by consensus. Study quality was assessed independently by two reviewers with the Joanna Briggs Institute (JBI) Critical Appraisal Checklist for Studies Reporting Prevalence Data, with higher scores indicating a lower risk of bias. The results are presented in Supplementary Table 1. Data from individual studies were extracted and entered into a standardized Excel spreadsheet designed for this study, including study and patient characteristics, country, year of infection, cohort size, disease severity, prevalence of AAN–IFN- I, survival outcomes, assay methods, age, sex distribution, vaccination status, and SARS-CoV-2 strain.

## Results

### Literature search

The PubMed search identified 187 articles. After the removal of duplicates and consolidation of the results across databases, we screened the titles and abstracts of the articles to identify original studies reporting primary data on the prevalence and/or functional characterization of AAN–IFN-I in patients with confirmed SARS-CoV-2 infection. Full texts were assessed for 62 studies(27, 33–39). In total, 54 studies were included in the narrative review(5, 7, 21, 40–89), and 20 studies were included in the meta-analysis on the basis of the presentation of functional data (neutralization assay)(5, 7, 21, 41, 45–47, 50, 54, 59, 60, 62, 68, 73, 74, 76, 80, 85, 88, 90). The study selection flowchart is shown in Figure 1.

**Figure 1:**
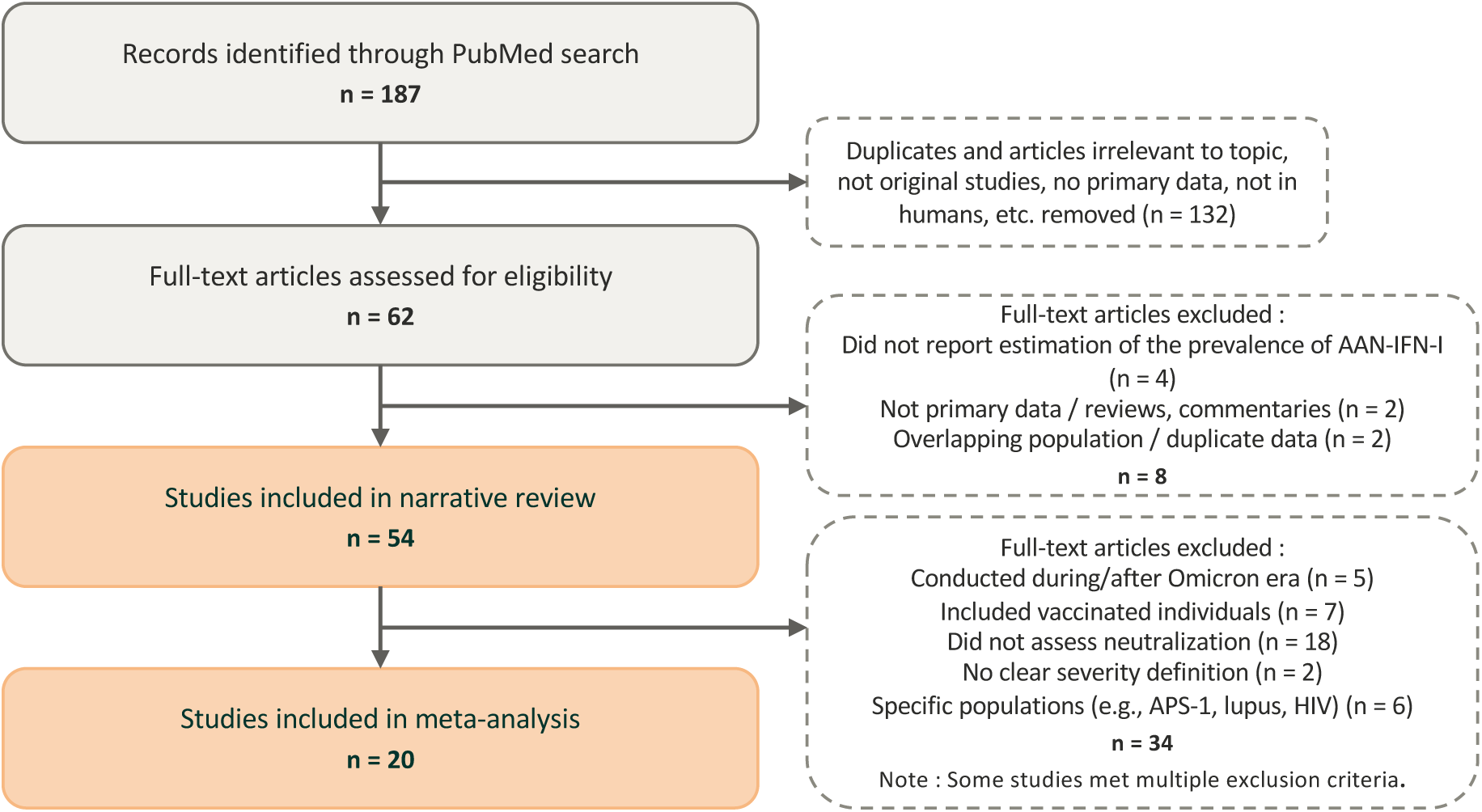
Flowchart showing study identification, screening, eligibility assessment, and selection for the systematic review and meta-analysis.

### Study characteristics

The 54 original studies included in the narrative review covered recruitment periods ranging from the first European wave of the pandemic to December 2022, with the seminal study recruiting patients between February and May 2020 (Figure 2). The studies included 65,687 individuals in total, including 19,545 patients with COVID-19 and 12,732 with severe or critical disease. The cohorts originated from Europe (Estonia, Denmark, France, the United Kingdom, the Netherlands, Italy, Spain, Germany, Sweden, Switzerland, Belgium, and Russia)(40, 41, 43, 45, 46, 48–50, 54, 59, 62–66, 68, 69, 71, 75, 75, 76, 78–82, 85, 87, 88), North America (United States)(47, 51, 56, 58, 61, 67, 72, 73, 83, 86), South America (Brazil, Colombia, and Peru)(44, 70, 84, 89), Asia (Japan and Singapore)(42, 60, 74), the Middle East (United Arab Emirates and Iran)(52, 53, 57), and North Africa (Morocco)(90). Five additional international studies included cohorts from 40, 38, 9, 7 and 7 countries, respectively(5, 7, 21, 55, 77). The worldwide geographic distribution of the cohorts included in this study is presented in Figure 3. The main characteristics of the studies included are described in Table 1, with additional information provided in Supplementary Table 2. Six studies focused on specific populations, such as patients with APS-1, HIV infection or systemic lupus erythematosus (SLE)(43, 69–71, 77, 81).

**Figure 2:**
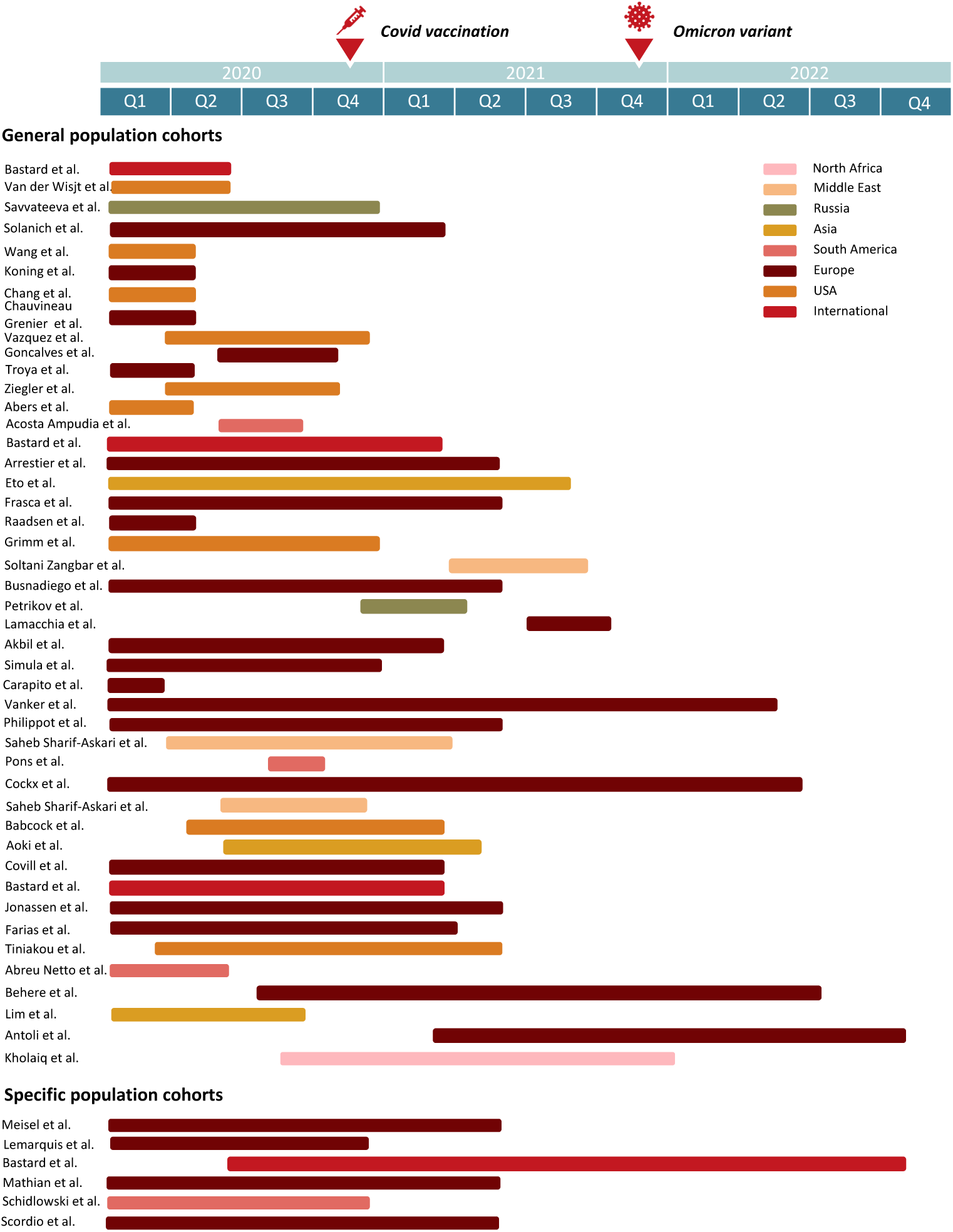
Recruitment periods of the studies included in the systematic review. Horizontal bars indicate the enrollment period of each study and are color-coded by geographic region. The timing of COVID-19 vaccination and the emergence of the Omicron variant are shown for reference.

**Figure 3:**
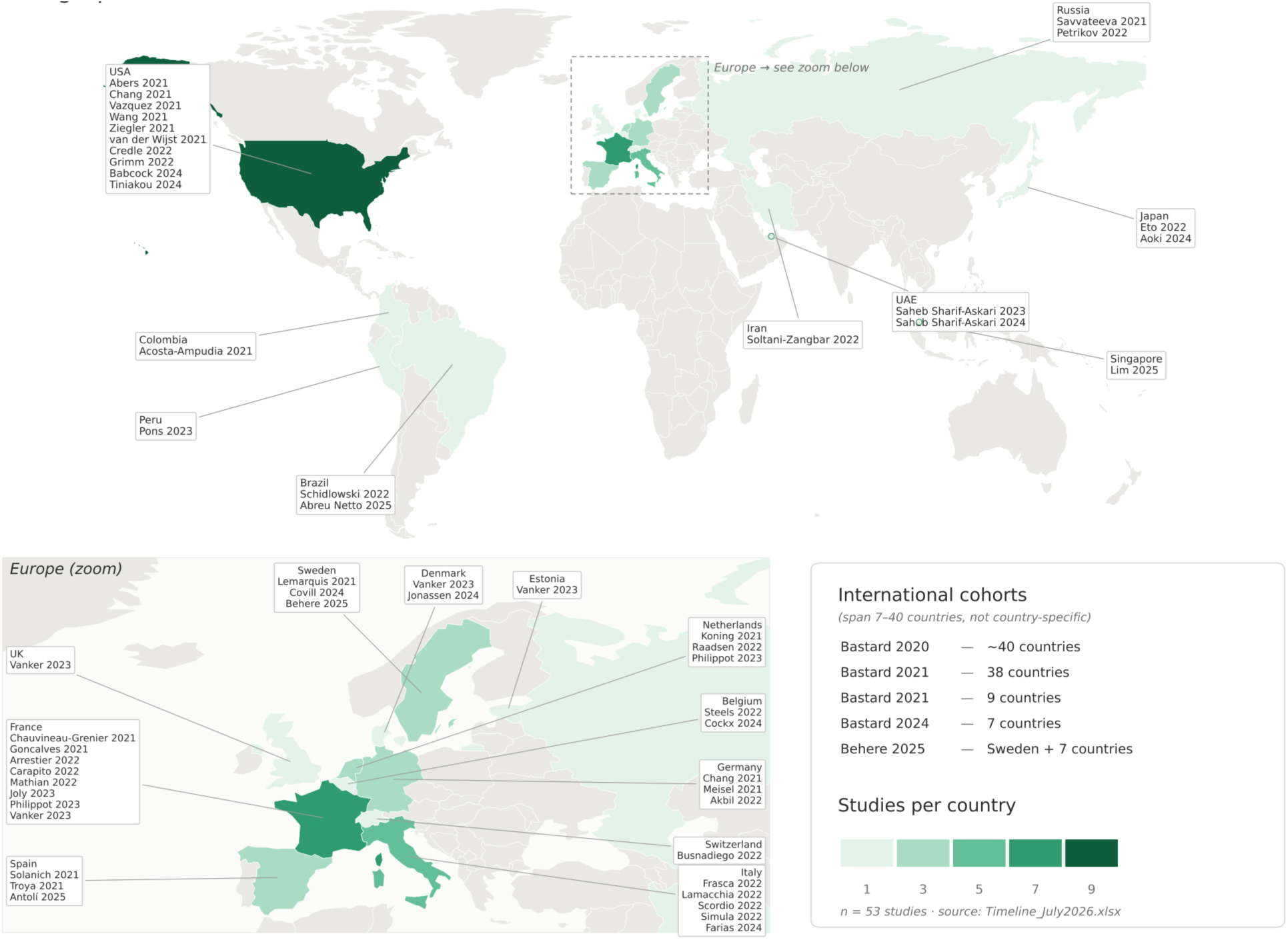
Geographic distribution of the studies included in the systematic review (First author, publication year). The intensity of the shading indicates the number of studies included. International multicenter cohorts spanning multiple countries are listed separately. *Map generated with the assistance of Claude (Anthropic)*.

**Table 1:**
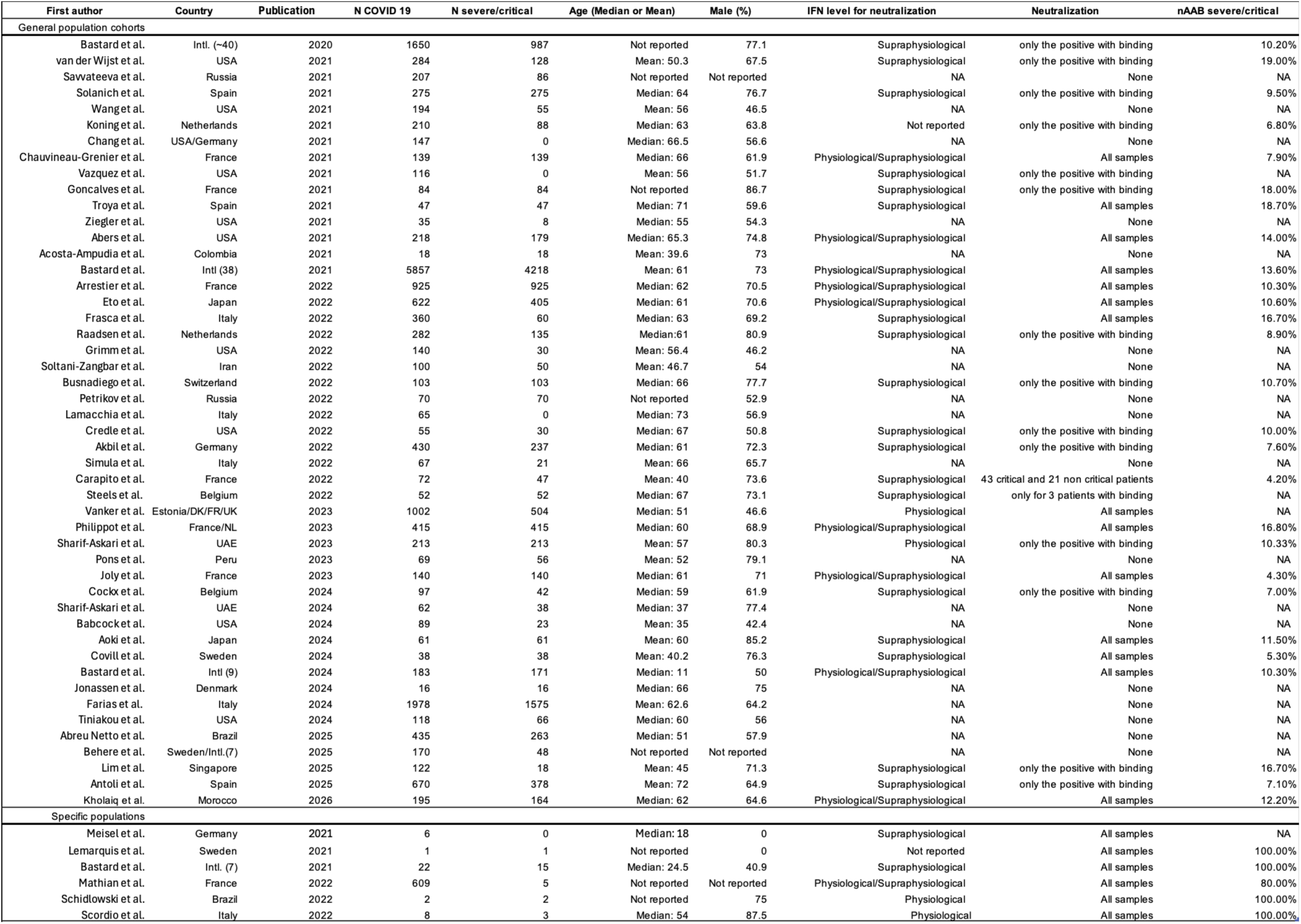
Characteristics of the studies included in the systematic review. For each study, the table shows the country, recruitment period, cohort size, age and sex distribution, COVID-19 severity, methods used for binding and neutralization assays, IFN-I subtypes tested, and the prevalence of binding and neutralizing anti–type I IFN autoantibodies. Abbreviations: AAB, autoantibodies; IFN-I, type I interferon; RT-PCR, reverse-transcription polymerase chain reaction; LIPS, luciferase immunoprecipitation system; RLBA, radioligand binding assay; NA, not applicable.

### Narrative review

#### (1) Binding assays versus functional neutralization assays

The studies included used two main approaches for the detection of auto-Abs against type I interferons (IFN-I): binding assays and functional neutralization assays. The binding assays for auto-Abs were based on ELISA, Gyros technology, or multiplex bead-based immunoassays and detected antibody binding without demonstrating the functional inhibition of IFN-I activity. By contrast, neutralizing activity was assessed in cell-based assays measuring the ability of patient serum to inhibit IFN-I–induced signaling or antiviral effects. Following the binding of IFN-I to IFNAR1/2, the activation of JAK1 and TYK2 induces STAT1/STAT2 phosphorylation, ISGF3 formation, and the transcription of interferon-stimulated genes involved in antiviral defense. Neutralizing auto-Abs disrupt this pathway upstream by binding IFN-I, thereby impairing IFN signaling, leading to weaker ISG induction, or a loss of IFN-mediated antiviral protection in infected cells (e.g., SARS-CoV-2 or VSV infection assays)(91).

Functional assessments of neutralization activity were performed in 36 of the 54 studies included: 14 performed such assessments only on samples yielding positive results in binding assays, 2 performed assessments on arbitrarily selected patients, and 20 performed functional assessments on all samples. The restriction of neutralization assays to samples testing positive in binding assays may result in an underestimation of the prevalence of functional neutralizing auto-Abs as low titers autoantibodies can be neutralizing(5). The remaining 18 studies assessed the prevalence of auto-Abs exclusively with binding assays. As binding assays do not demonstrate the functional impairment of type I IFN immunity, we restricted the meta-analysis to studies assessing neutralizing activity.

#### (2) IFN concentrations used for neutralization assays

Very different IFN concentrations were used for the neutralization assays, ranging from physiological concentrations (∼100 pg/mL) to supraphysiological concentrations (∼10 ng/mL). As the circulating IFN-I levels measured *in vivo* are substantially lower, the use of physiological concentrations probably provides a more accurate estimate of biologically relevant neutralizing activity. By contrast, the use of higher IFN concentrations may lead to an underestimation of the proportion of patients carrying functionally relevant AAN–IFN-I and their impact on susceptibility to viral infections. Four of the studies included used near-physiological IFN concentrations only, 20 used supraphysiological concentrations only, and 10 assessed both. IFN concentrations were not specified in two studies.

#### (3) Type of IFN evaluated

Functional assays investigated primarily auto-Abs neutralizing IFN-α and IFN-ω, the two type I interferons for which impaired activity or deficiency is most consistently associated with severe COVID-19. A smaller number of studies evaluated auto-Abs against IFN-β. In total, 54 studies assessed the neutralization of IFN-α, 39 studied the neutralization of IFN-ω, and 23, the neutralization of IFN-β. Differences in the IFN subtype evaluated probably contributed to between-study heterogeneity in prevalence estimates.

#### (4) Categories of patients included

The patient populations included also differed considerably between studies. Cohorts variably included asymptomatic or mild outpatient cases, hospitalized patients with pneumonia, severe/critical ICU patients, patients who died, pediatric cohorts, or selected populations with underlying autoimmune or genetic disorders, such as APS-1 or SLE. The definitions of disease severity and inclusion criteria also differed between studies.

### Specific backgrounds

Six studies focused on specific backgrounds, including APS-1 (*n*=4)(69–71, 77), HIV infection (*n*=1)(81), and SLE (*n*=1)(43); all six assessed neutralizing activity. In APS-1 cohorts, auto-Abs neutralizing IFN-α and IFN-ω were detected in all infected individuals, including 22/22 patients in the international cohort (15 severe/critical cases, 4 deaths), 4/4 infected patients in the German cohort (all with mild disease), 2/2 brothers with life-threatening COVID-19 in the Brazilian study, and 1/1 child with severe disease in the Swedish study. In the HIV cohort, 7 of 8 patients with COVID-19 carried AAN–IFN-I, including all three patients with severe or critical disease. Two patients died, one from COVID-19 and one from cerebral non-Hodgkin lymphoma; both carried AAN–IFN-I. For the SLE study, 609 patients with COVID-19 were included, five of whom had severe or critical disease, including four carrying AAN–IFN-I. These findings support the notion that AAN–IFN-I can result from defects of immune tolerance, including, in particular, the genetic etiology underlying APS-1, and that, in such cases, they are associated with a high risk of critical COVID-19 across age groups. More generally, autoimmunity should raise concerns about the possible presence of such auto-Abs and the risk of severe or critical disease.

### Pooled prevalence of AAN-IFN-I in COVID-19 patients

Twenty studies on unvaccinated individuals from the general population in which severity criteria were defined were included in the meta-analysis(5, 7, 21, 41, 45–47, 50, 54, 59, 60, 62, 68, 73, 74, 76, 80, 85, 88, 90): 12 European cohorts, 3 international cohorts, 2 Asian cohorts, 2 North American cohorts, and 1 North African cohort. These studies comprised a total of 52,187 individuals, including 11,380 patients with COVID-19 and 7,814 with severe or critical disease, and 3,318 with moderate, mild or asymptomatic disease. All these studies assessed the prevalence of AAN–IFN-I with functional neutralization assays; in seven of these studies, neutralization assays were performed exclusively on samples testing positive in binding assays. We first estimated the overall prevalence of neutralizing AAN–IFN-I across the 19 studies. The pooled prevalence was 7.2% (95% CI, 5.6–9.1) (Figure 4), with substantial between-study heterogeneity (I² = 88.3%). We investigated potential sources of between-study heterogeneity by performing univariable study-level meta-regression analyses to assess the association of pooled prevalence with neutralization assay strategy (all samples vs. binding-positive samples only), median age, the proportion of male subjects, the proportion of severe/critical cases and region of the world. Only the proportion of severe/critical cases was significantly associated with the pooled prevalence estimate, with higher estimate for studies with higher proportions of severe/critical cases (β (SE) = 1.26 (0.38), *p*=0.001) (Table 2).

**Figure 4:**
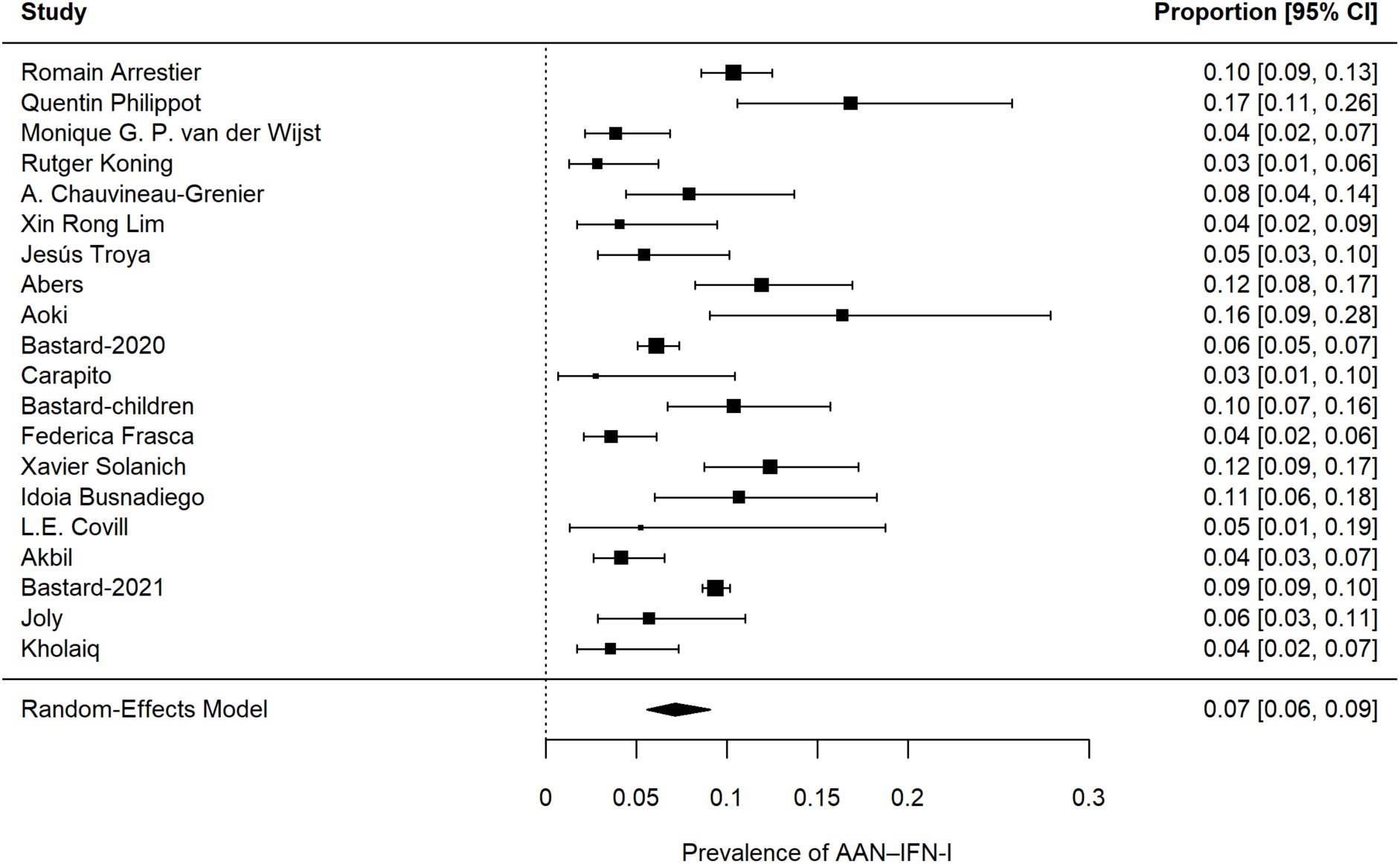
Forest-plot of the pooled prevalence of AAN-IFN-I.

**Table 2:**
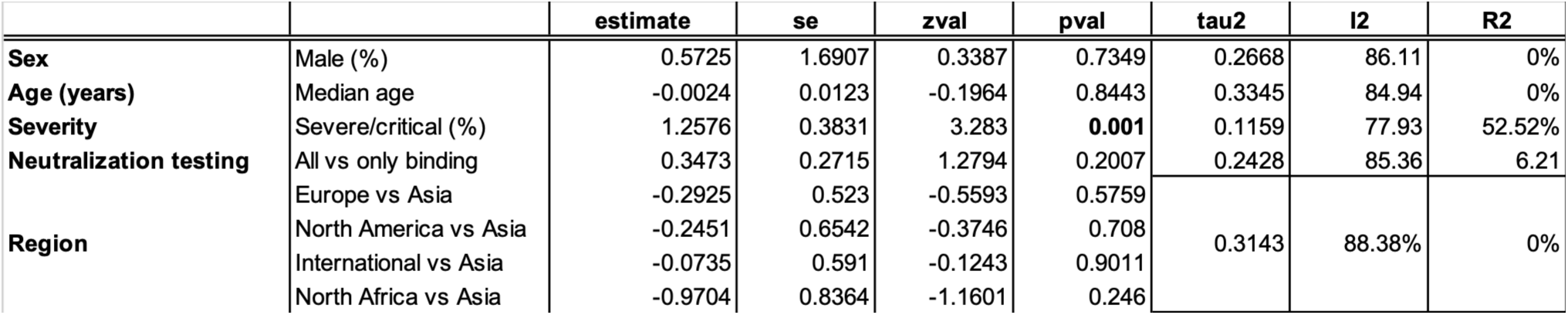
Univariable random-effects meta-regression of study-level factors associated with the prevalence of neutralizing anti-type I interferon autoantibodies (AAN–IFN-I). Estimates, standard errors (SE), *z* values, *P* values, residual heterogeneity (τ² and I²), and the proportion of heterogeneity explained (R²) are shown.

### Pooled prevalence of AAN-IFN-I by COVID-19 severity

We compared the estimated prevalence of AAN-IFN-I between the severe/critical and mild/moderate COVID-19 groups. In a subgroup meta-regression, COVID-19 severity was strongly associated with the prevalence of AAN–IFN-I (OR [95%CI] = 11.9 [7.7 – 18.4], *p* = 5×10^-29^). Disease severity accounted for almost all the between-study heterogeneity (R² = 97.8%), with only a low level of residual heterogeneity (I^2^ = 27%). Sensitivity analyses excluding the largest study(5) yielded consistent estimates for the association between severity and prevalence (OR [95%CI] = 11.4 [6.0 – 21.7], *p*-value = 10^-13^) and further reduced residual heterogeneity (I² = 13.9%), confirming the robustness of the findings. We then used random-effects models to estimate the pooled prevalence separately within each severity stratum. The pooled prevalence of AAN–IFN-I in patients with asymptomatic/mild/moderate disease was 1.0% (24/3318; 95% CI, 0.7 - 1.5%; Figure 5), with negligible between-study heterogeneity (I² = 0%). The exclusion of moderate cases yielded a similar prevalence estimate of 0.9% (17/2649; 95% CI, 0.6% - 1.4%). For physiological doses of type I IFN, the prevalence of AAN-IFN-I in patients with asymptomatic/mild disease was significantly lower than that in the general population (1.0% vs. 2.2%, *p* = 0.0025, Supplementary Table 3). The pooled prevalence of AAN–IFN-I in patients with severe/critical COVID-19 was 11.2% (909/7,814; 95% CI, 9.9 – 12.5%), with study-specific prevalence estimates ranging from 4.3% to 19.1% across the 20 studies included, and moderate between-study heterogeneity (I² = 39.2%). If the analysis was restricted to auto-Abs neutralizing supraphysiological doses of type I IFN, the pooled prevalence in patients with severe/critical disease was estimated at 6.3% (420/5405; 95%CI, 4.7 – 8.5%), with between-study heterogeneity was high (I² = 72.4%). In deceased patients, the estimated pooled prevalence of AAN–IFN-I was higher, at 16.0% (311/1,871; 95% CI, 12.9–19.8%), with moderate between-study heterogeneity (I² = 43.1%).

**Figure 5:**
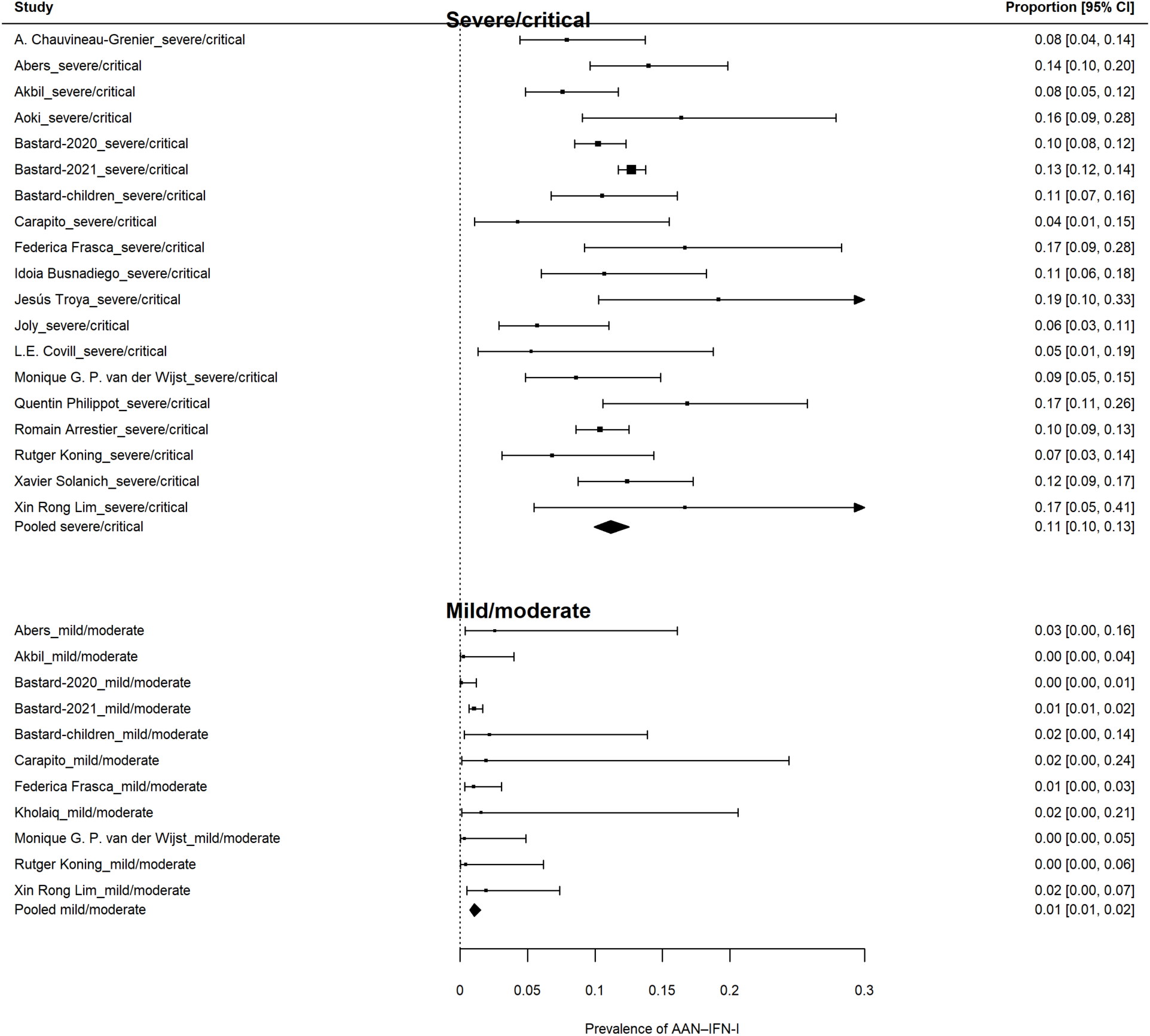
Forest plot of the pooled prevalence of AAN-IFN-I by COVID-19 severity.

#### Effect of age on the prevalence of AAN-IFN-in patients with severe/critical disease

We then refined the analysis by evaluating the association of age with the estimated pooled prevalence of AAN–IFN-I in patients with severe/critical disease. A subgroup meta-regression showed that age was significantly associated with the prevalence of AAN–IFN-I (OR [95%CI] for >65 years old vs. <65 years old = 1.71 (1.12 – 2.59), *p* = 0.01), although between-study heterogeneity was high (I²=77.9%). Similar results were obtained when the largest study(5) was excluded. The estimated pooled prevalence of AAN-IFN-I was 6.3% (456/5150; 95% CI, 4.5 – 8.7) in patients under the age of 65 years and 11.2% (431/3413; 95% CI, 9.2 – 13.5) in patients over the age of 65 years (Figure 6). The prevalence of these autoantibodies was higher in patients with severe/critical COVID-19 than in the general population, for both age groups (1.6% before the age of 65 years and 4.8% thereafter).

**Figure 6.**
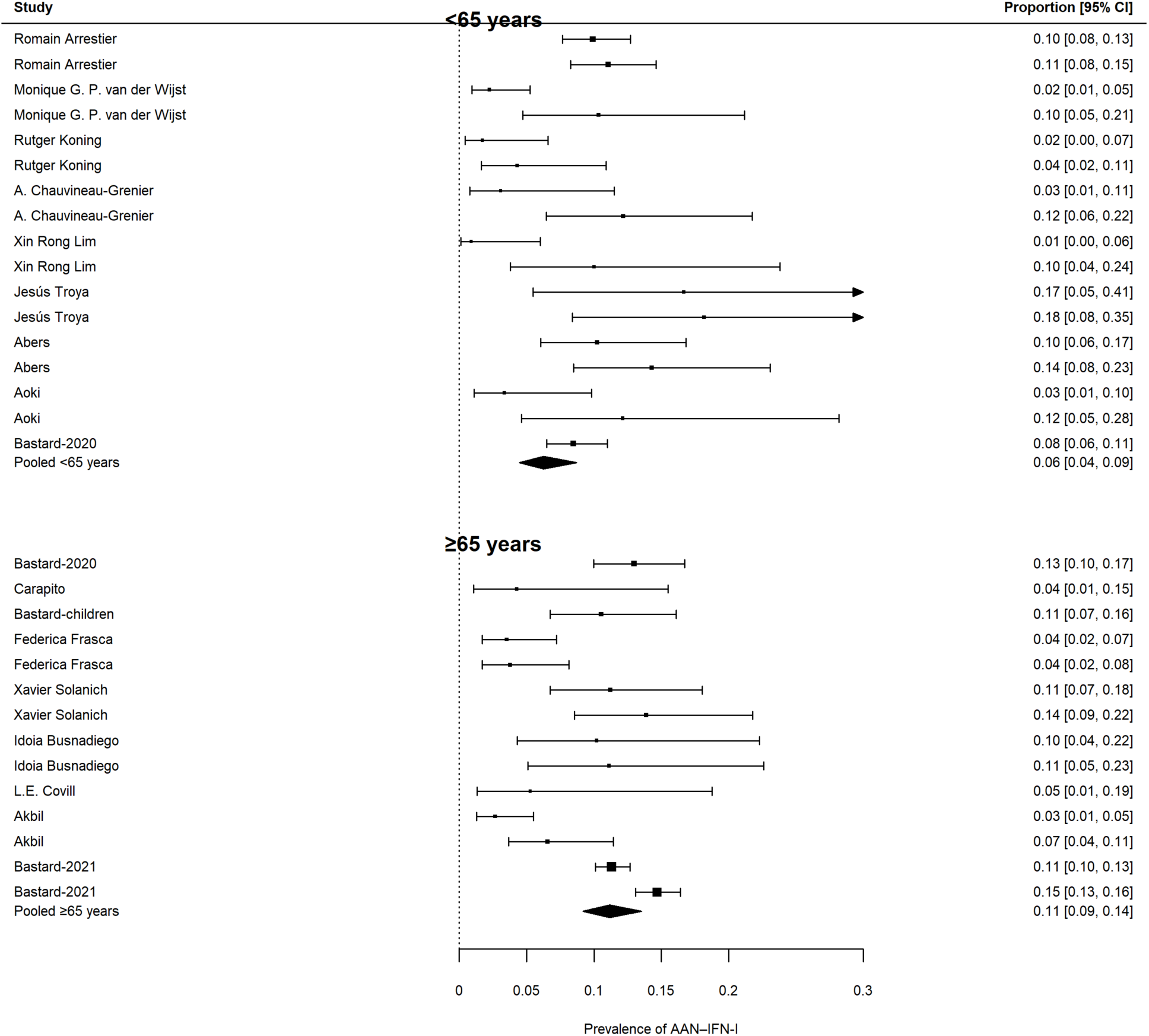
Forest plot of the pooled prevalence of AAN-IFN-I in patients with severe/critical COVID-19, by age.

## DISCUSSION

We show that AAN-IFN-I have consistently been detected in independent cohorts worldwide, with the pooled prevalence of AAN–IFN-I estimated at 7.9% (95% CI, 6.0–10.4) across all severity groups, increasing to 11.4% (95% CI, 10.2–12.7) in patients with severe or critical COVID-19 and 15.3% (95% CI, 12.1–19.2) in patients who died. Disease severity was by far the strongest determinant of AAN–IFN-I prevalence across studies, accounting for almost all the between-study heterogeneity of pooled prevalence. No significant difference was found between regions of the world for the prevalence of AAN–IFN-I in patients with severe or critical disease. The prevalence of AAN-IFN-I was significantly higher in patients with life-threatening COVID-19 over the age of 65 years. This age-related distribution is consistent with previous studies showing that rare inborn errors of type I IFN immunity account for a larger fraction of life-threatening COVID-19 in younger patients and highlights the substantial contribution of AAN-IFN-I to the global burden of severe SARS-CoV-2 infection(6, 20). Together, these findings establish that defective type I IFN immunity, whether caused by AAN–IFN-I or by rare IEIs affecting the IFN-I pathway, constitutes a major mechanism underlying life-threatening COVID-19 in geographically diverse populations.

There is growing evidence that AAN–IFN-I confer a predisposition to severe disease due to multiple other viruses in addition to SARS-CoV2(92). They are detected in about 5% of patients with life-threatening influenza pneumonia, particularly those under the age of 70 years(93), and in about one quarter of patients hospitalized with Middle East respiratory syndrome(94). An even greater enrichment is observed for arboviral infections, with AAN-IFN-I detected in about 10% of severe tick-borne encephalitis and up to 40% of West Nile virus encephalitis cases in international cohorts(95). They have also been identified in the most severe cases of Powassan, Usutu, and Ross River virus infections reported to date(96). In addition, AAN–IFN-I account for approximately one third of life-threatening adverse reactions to vaccination against yellow fever with the live attenuated viral vaccine and have recently been identified in severe adverse reactions to yellow fever/chikungunya vaccination(97, 98). Finally, AAN-IFN-I have also been detected in two lethal cases of avian influenza(99). Collectively, these observations establish AAN–IFN-I as a determinant of susceptibility to common to a number of severe viral diseases and raise questions as to their origin.

The mechanisms leading to AAN–IFN-I production remain incompletely understood, but studies of several IEI and thymic disorders have suggested a key role for defects of thymic function and immune tolerance, including APS-1 due to AIRE deficiency, IPEX syndrome caused by *FOXP3* mutations, and combined T- and B-cell immunodeficiencies associated with hypomorphic *RAG1* or *RAG2* mutations(8, 93, 100, 101). The high prevalence of neutralizing AAN–IFN-I in individuals with APS-1, HIV infection, or SLE provides further support for the concept that impaired immune tolerance(43, 69–71, 77, 81), whether monogenic or otherwise, creates a predisposition for the development of these antibodies. Future studies should define the genetic and immunological determinants of AAN–IFN-I more precisely, to guide indications for systematic screening in the general population, in patients with severe viral infections, and before the administration of live-attenuated viral vaccines, and perhaps in the uninfected elderly population to prevent future diseases.

## Data Availability

All the data included in this study are available from the corresponding author upon request.

## Contributors

JLC, QZ, PB, AC conceived the study.

JLC, QZ, PB, AC, EF developed the review protocol.

QZ, PB, EF independently screened the studies.

EF extracted data and conducted the literature search.

AC performed the meta-analysis and statistical analyses.

EF drafted the manuscript; all the authors provided critical input during the writing of the manuscript.

The corresponding author attests that all the listed authors meet the authorship criteria and that no others meeting the criteria have been omitted.

## Ethics approval

Not required.

## Data sharing

All the data included in this study are available from the corresponding author upon request.

## Acknowledgments

We thank the authors of the studies included in this systematic review who provided additional data or study-specific information upon request. We thank all members of the Laboratory of Human Genetics of Infectious Diseases in Paris and Dallas. Figure 3 (world map) was generated with the assistance of Claude (Anthropic).

---

**Supplementary Table 1:**
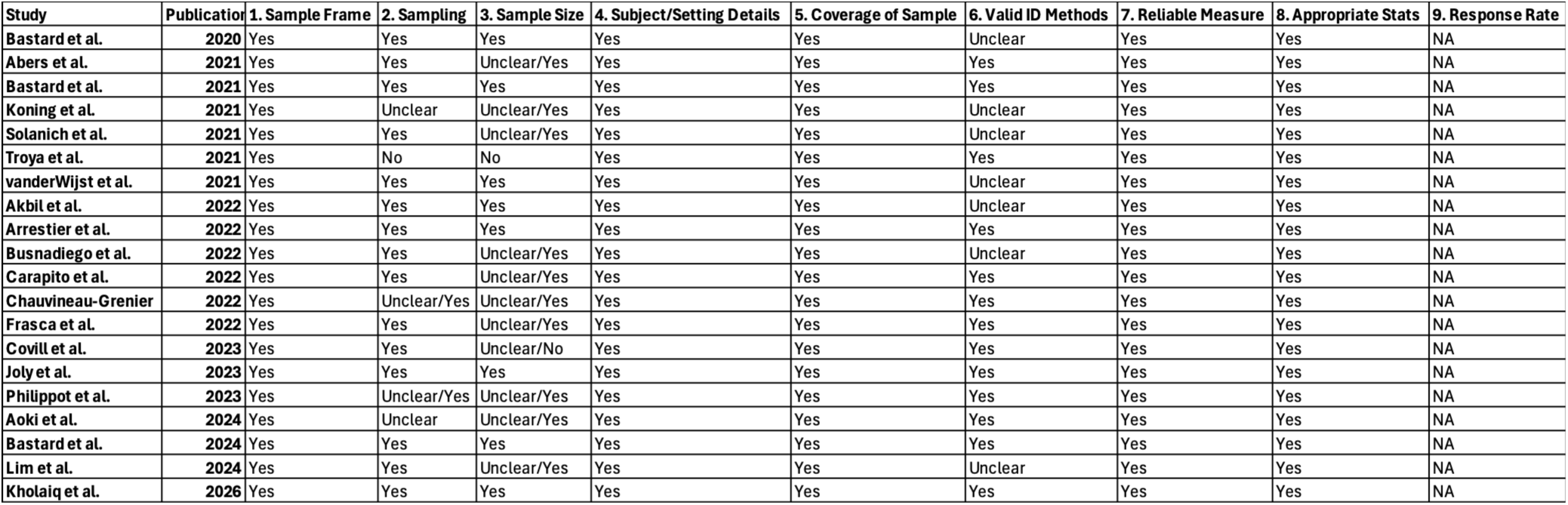
Assessment of the quality of the studies included in the systematic review according to the Joanna Briggs Institute (JBI) Critical Appraisal Checklist for Studies Reporting Prevalence Data. Study quality was assessed independently by two reviewers. Higher scores indicate a lower risk of bias.

**Supplementary Table 2.**
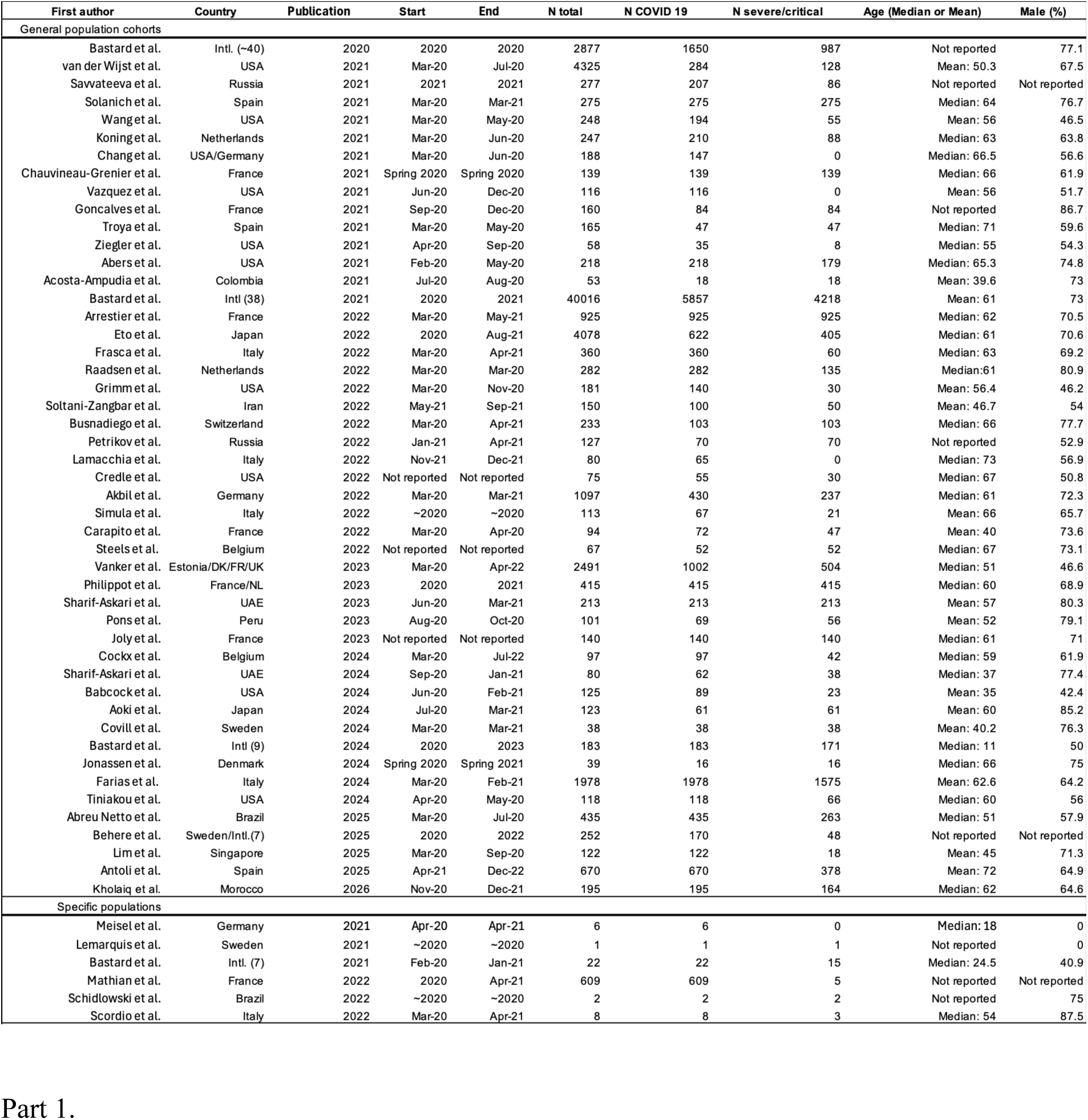
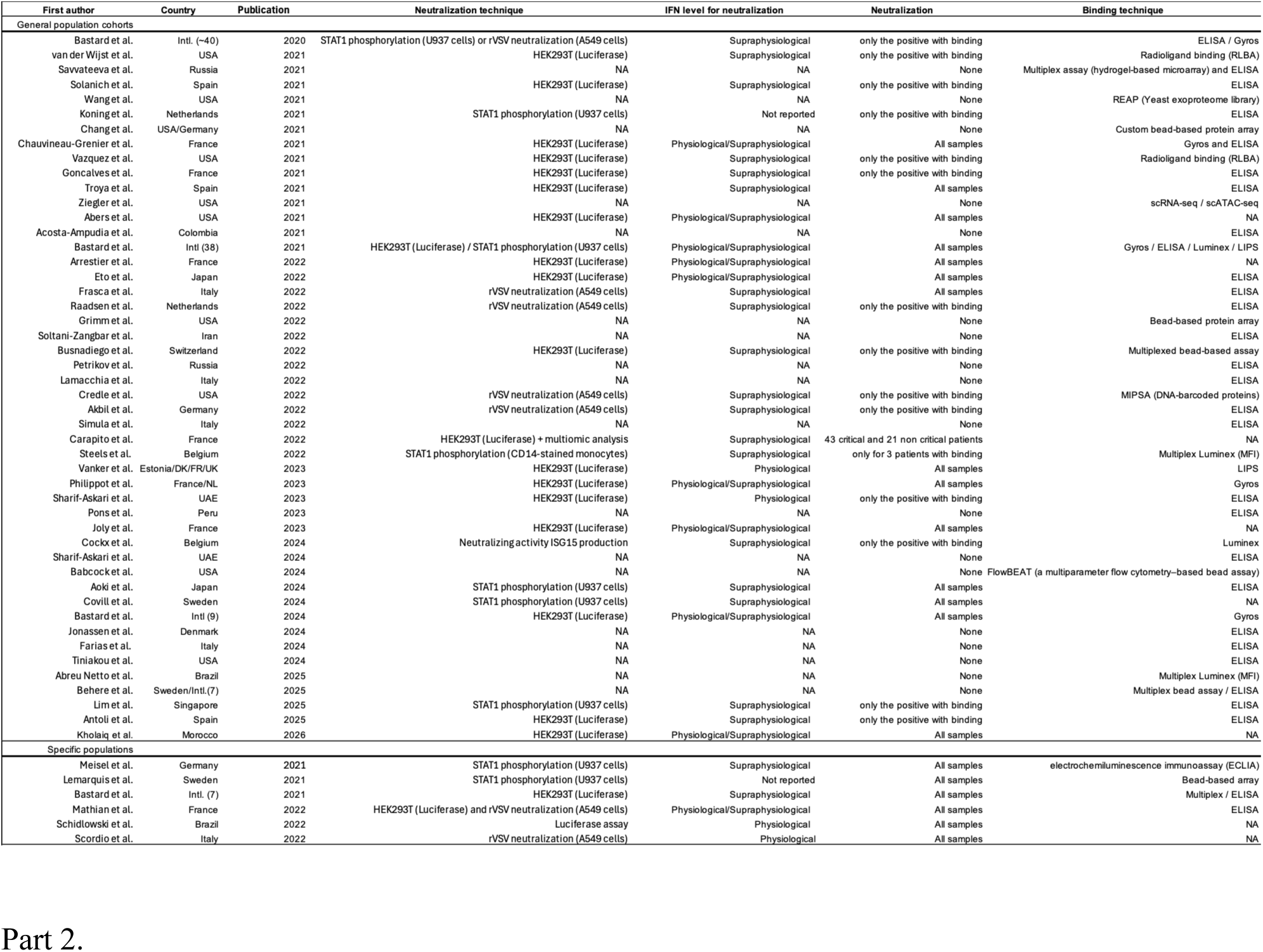
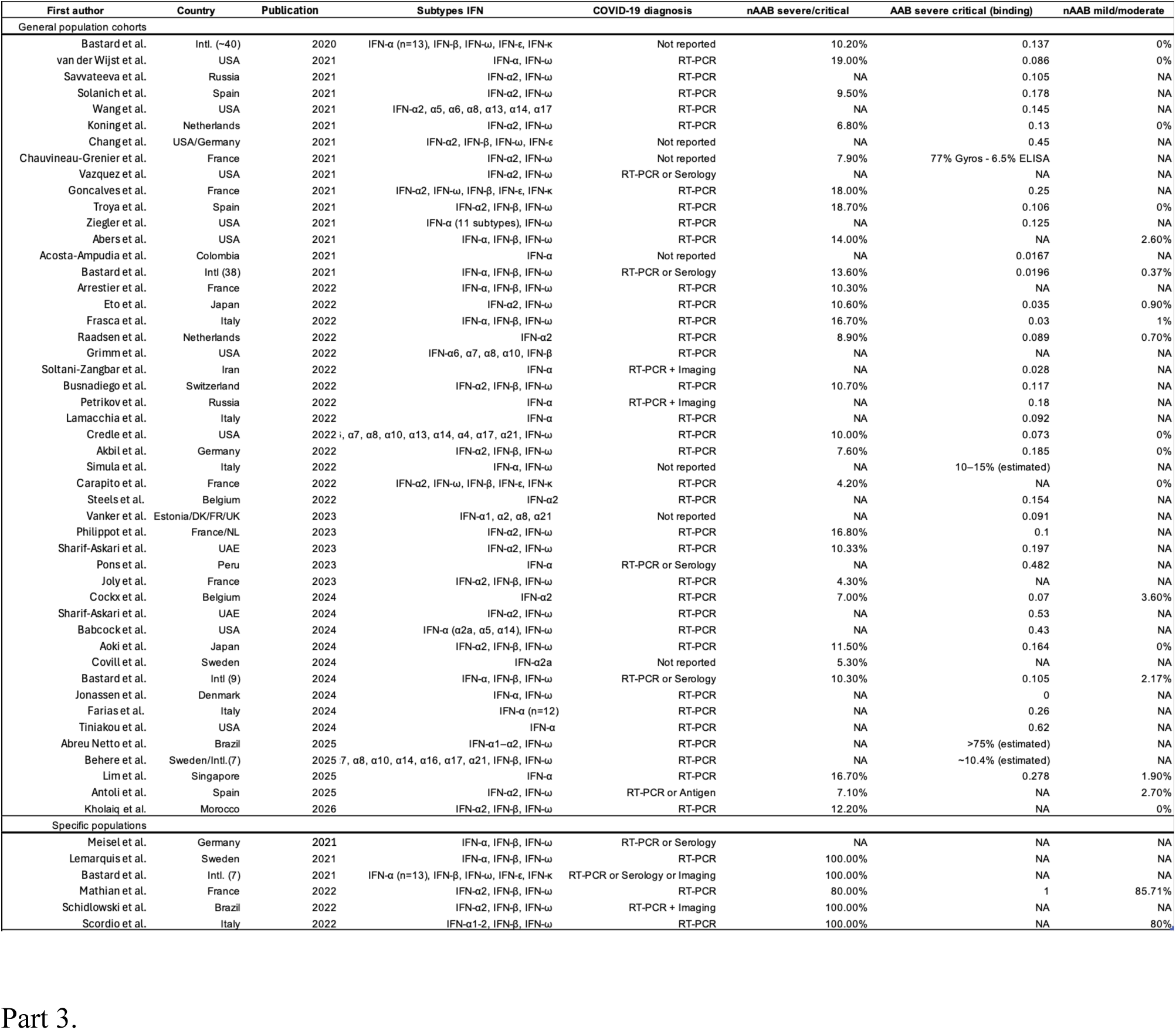
(Parts 1–3): Complete characteristics of the studies included in the systematic review and meta-analysis.

**Supplementary Table 3:**
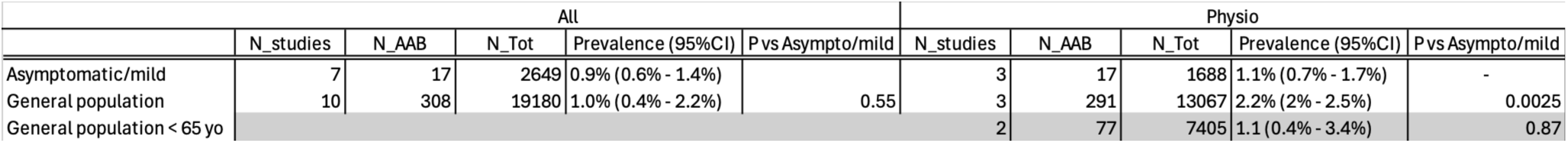
Prevalence of autoantibodies neutralizing type I interferon in patients with asymptomatic/mild COVID-19 disease and the general population. Pooled prevalence estimates are shown for all studies and for studies assessing neutralization at physiological concentrations of interferon. *P* values are provided for comparisons of prevalence between the general population and patients with asymptomatic/mild COVID-19.

